# A Lagrangian Approach Towards Quantitative Analysis Of Flow-mediated Infection Transmission In Indoor Spaces With Application To SARS-COV-2

**DOI:** 10.1101/2021.08.22.21262447

**Authors:** Joseph Wilson, Shelly L. Miller, Debanjan Mukherjee

**Affiliations:** Department of Mechanical Engineering, University of Colorado Boulder

**Keywords:** Lagrangian, Respiratory Infection, Finite Time Lyapunov Exponents, Particle Transport, Airborne Transmission

## Abstract

The ongoing SARS-CoV-2 (Covid-19) pandemic has ushered an unforeseen level of global health and economic burden. As a respiratory infection, Covid-19 is known to have a dominant airborne transmission modality, wherein fluid flow plays a central role. Quantification of complex non-intuitive dynamics and transport of pathogen laden respiratory particles in indoor flows has been of specific interest. Here we present a Lagrangian computational approach towards quantification of human-to-human exposure quantifiers, and identification of pathways by which flow organizes transmission. We develop a Lagrangian viral exposure index in a parametric form, accounting for key parameters such as building and layout, ventilation, occupancy, biological variables. We also employ a Lagrangian computation of the Finite Time Lyapunov Exponent field to identify hidden patterns of transport. A systematic parametric study comprising a set of 120 simulations, yielding a total of 1,320 different exposure index computations are presented. Results from these simulations enable: (a) understanding the otherwise hidden ways in which air flow organizes the long-range transport of such particles; and (b) translating the micro-particle transport data into a quantifier for understanding infection exposure risks.

## 1 Introduction

Respiratory infection outbreaks have been a major public health concern in recent times, with perhaps the most massive global burden observed due to the SARS-CoV-2 (Covid-19) outbreak. At the time of writing this article, more than 200 million cases and over 4.2 million deaths have been reported worldwide [1]. This has been accompanied by widespread global economic impacts due to lockdown and closure of schools, offices, and businesses [34]. In addition to Covid-19, recent decades have also seen several other infection outbreaks of concern, such as SARS, MERS, and H1N1 influenza [23,42,15]. The public health impacts of these disease outbreaks have placed a renewed emphasis on comprehensive understanding of infection transmission and control measures. A wide range of existing studies, many of which have been comprehensively summarised in recent surveys [47], collectively point towards several key aspects regarding infection transmission risks for respiratory infections. First, operations in occupied indoor spaces pose specific challenges owing to the higher potential of infection spread as substantiated in multiple studies - examples include restaurants [33]; hospitals and nursing homes [35,36,43]; call centers [45]; and musical choir practice halls [37] amongst others. The infection transmission risks are intimately connected to factors like ventilation and occupancy density [30]. Second, there is substantial evidence indicating that transmission of infections like SARS-CoV-2 through respiratory ejecta particles has a dominant air-borne modality [29,39,24]. Infection spread is increasingly acknowledged to occur via a *“short-range route”*, where larger respiratory particles or droplets are likely the main agent of transmission; and a *“long-range route”* where aerosolized small scale or micro-particles are likely the main agent of transmission [28,59]. Third, a growing body of evidence indicates that SARS-CoV-2 transmission dynamics is a multiscale problem with an intimate flow physics connection [38,10]. The infection transmission process comprises micro-scale phenomena involving viral particles and droplets, their generation and their behavior; intermediate or meso-scale host-to-host transmission events; and macro-scale dynamics of infection spread and recovery at a community or population level. Fluid mechanics plays a central role at each of these scales [38,11]. Complex flow phenomena underlie the generation of respiratory droplets during coughing, sneezing, and exhalation [12,50]. Fluid mechanics of forceful exhalation spreads the respiratory ejecta [17], which are further dispersed by background airflow and turbulence [19]. Individual droplets or particles behave under the action of forces from background flow, gravity, and evaporation, causing them to settle down or stay suspended based on their size [61,44,57,14]. Background air flow in indoor spaces are in turn related to ventilation operations [30,40]. Together, these three aspects inform any attempts towards quantifying host-to-host infection transmission risks.

Owing to the important role played by fluid flow, several approaches to account for flow phenomena in infection transmission within enclosed spaces (*and otherwise*) have been outlined. The classical Well’s evaporation-falling curve for respiratory droplets is amongst the earliest of such works [60,61], which has found some renewed attention amidst the Covid-19 pandemic [16]. Models based on the Wells-Riley equation [49,41] (*originally devised for airborne measles transmission*) are widely used for probabilistic infection transmission risk estimations in indoor ventilated spaces, employing the concept of a ‘quanta’ of infection. Dose-response based models are an alternative approach, based on the concept of estimating impact of exposure to a certain dose of the infecting agent [54]. Both of these statistical models are based on several assumptions, including that of well-mixed homogenized systems - an aspect that has been further investigated in recent studies on indoor airborne Covid-19 infection risks [8]. This assumption coarse-grains the finer scale flow information in indoor spaces. Such level of detail is, however, the focus of larger-scale computational fluid dynamics (CFD) models. Several attempts at using CFD-based approaches have been outlined for indoor infection transmission analysis [58,32] - including advection-diffusion models for infectious particles [27], combined Eulerian and dose-response models [18], and tracer-gas simulations [21,26]. The majority of such attempts involve grid-based Eulerian approaches using space-time varying continuum fields. Despite the computational advantages of Eulerian approaches, many aspects of the underlying transport processes need an inherent physical description of droplets and micro-particles which are essentially Lagrangian quantities.

Here, we develop a Lagrangian approach for analysis of human-to-human transmission of infection in indoor spaces. Our focus in this study is to explore the heterogeneities in transport of respiratory micro-particles. Such aerosolized particles remain suspended for long durations, thereby undergoing complex non-intuitive spatiotemporal dynamics en route from one human to another. We seek to address two specific objectives: (a) understand the otherwise hidden ways in which air flow organizes long-range transport of such particles; and (b) translate the micro-particle transport data into a quantifier for understanding infection exposure risks. For the latter we develop a Lagrangian viral exposure index, and set up the framework for quantifying this index as a function of key parameters. For the former, we employ a Lagrangian computation of Finite Time Lyapunov Exponent fields to illuminate flow-mediated organization of particle transport. Thus, our goal is not to explore in-depth aspects of specific flow physics modeling intricacies, but to develop a methodology that connects indoor space flow modeling with exposure and transmission risks within a single unified framework. Here, we demonstrate these aspects through a parametric case-study for SARS-CoV-2 microparticles.

## 2 Methods

### 2.1 Framework and Study Design

Here we describe the proposed framework for analysis of respiratory ejecta particle transport and computing a viral exposure index (*using the notation V E*_*L*_) from a Lagrangian perspective. In a mathematically generalized form we describe this index as a function of four sets of parameters (*denoted by* **<u>Λ</u>**). We denote building layout, floor-plan, and dimension parameters in the parameter vector **<u>Λ</u>**_*b*_. Parameters related to ventilation including vent layouts and dimensions, flow rates, ACH etc. are denoted in the parameter vector **<u>Λ</u>**_*v*_. Occupancy pattern is denoted by the vector **<u>Λ</u>**_*o*_. Finally, respiratory particle details, including virus/pathogen parameters are denoted in the vector **<u>Λ</u>**_*p*_. Furthermore, assuming that floor-plan or dimension data can be described in form of a coordinate system associated with the layout, the occupancy pattern can be outlined in terms of *N*_*o*_ the number of occupants, and ***<u>X</u>***_*k*_ the location of each individual occupant, such that:

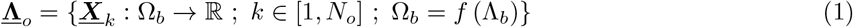

We propose to compute a Lagrangian Viral Exposure Index (*V E*_*L*_) as a function of the aforementioned parameter spaces:

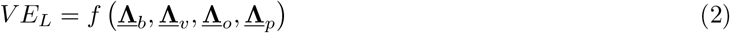

Using this overall mathematical framework, we outline here a simulation based case-study to elucidate the computation of the proposed *V E*_*L*_ index and its interpretation in the context of flow physics.

For our case-study, we select a simple model of a single room - dimensions 4m × 4m × 3m; with a single door (*closed*), a single window (*effective air ventilation area of 0*.*03125 m*^*2*^); and no furniture or other space modifications. The room size leads to around 173 sq.ft. floor area, which is used in commercial real estate estimates as equivalent to a small office space or meeting room. The room was supplied with one inflow and one exhaust vent of dimensions 0.3m × 0.15m × 0.3m. Ventilation was characterized using Air Changes per Hour or ACH, a measure of the number of times the air within a defined space is replaced in one hour. The ventilation flow rate was adjusted to achieve an ACH of 3.0, which is in the range of American Society of Heating, Refrigerating, and Air-conditioning Engineers (ASHRAE) recommendation for occupied public offices. Six different combinations of inflow and exhaust vent layouts were modeled by only changing the locations of the vents (*see Figure 1*). Three of them have inlet and exhaust located on opposite walls (*models: 180UU, 180UL, 180LL respectively in Figure 1*), and the remaining have inlet and exhaust located on perpendicular walls (*models: 90UU, 90UL, and 90LL respectively in Figure 1*). Within the room, five different infected hosts or index patients were considered, and occupancy patterns **<u>Λ</u>**_*o*_ were imposed for other non-infected subjects for whom we estimated *V E*_*L*_. Varying levels of respiratory particle transport were considered by varying effective particle diffusivity in flow.

**Figure 1:**
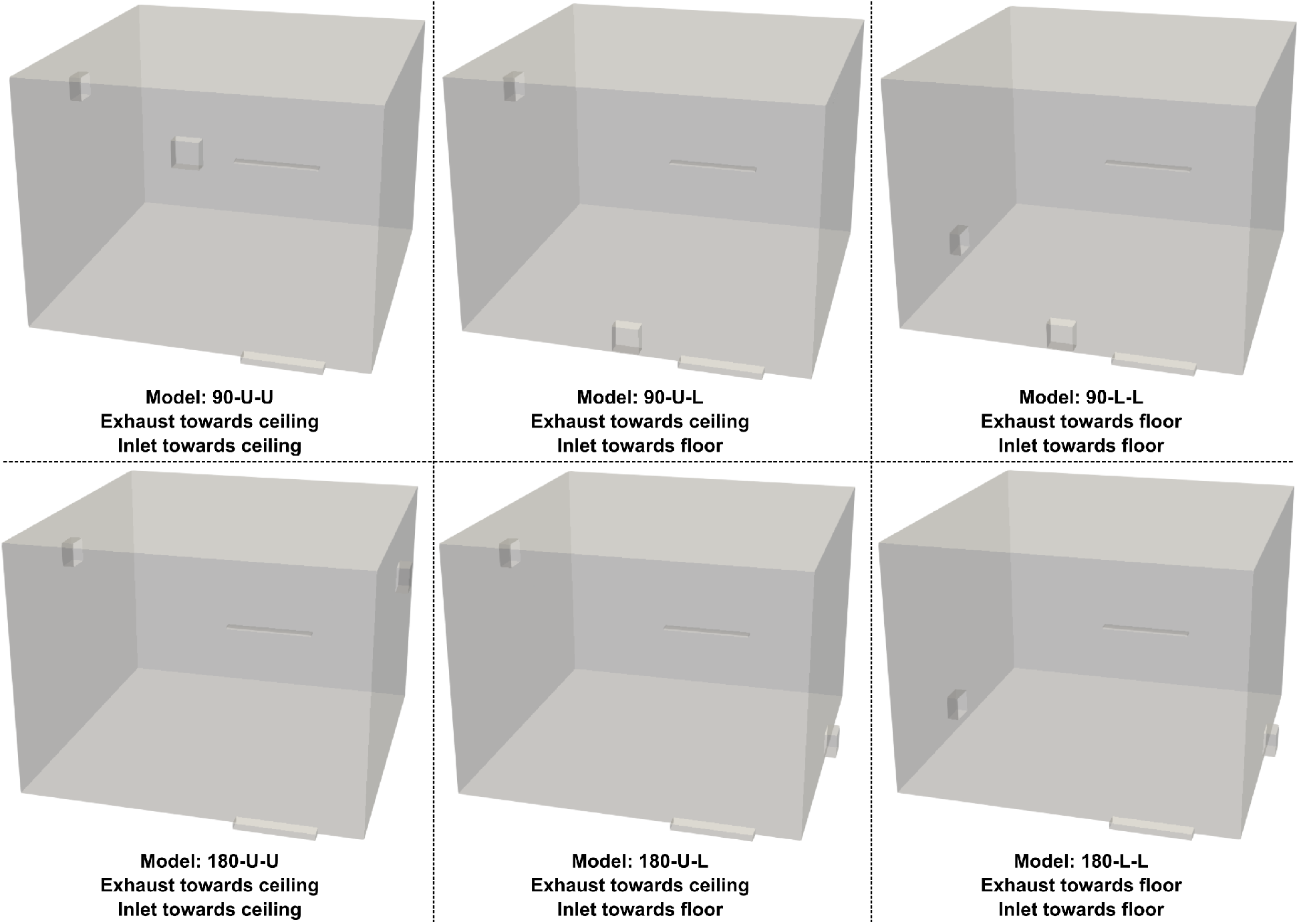
Illustration of the six different room ventilation configurations (**<u>Λ</u>**_v_) employed in the numerical case-study.

Considering four cases of particle dynamics, with five infected hosts, and six vent layouts, a total of 120 systematic simulations were conducted to investigate parametric variations in computed *V E*_*L*_ and subsequent predictions of infection transmission risk and its relation with indoor flow. For all simulations in the case-study we used steady Reynolds Averaged Navier Stokes (RANS) model for airflow with boundary conditions derived from air handling operations, as described in Section 2.3. This is an approach that has been validated against measured on-site data in a previous study of ours involving airflow in an entire isolation ward of a skilled nursing facility [36]. While RANS provides less resolved flow description than Large Eddy Simulations (LES), the lower computational expense of RANS models enabled pursuing the large set of parametric simulations. Airborne respiratory particles are assumed to be in the low-inertia tracer limit as micro-particles, and handled using one-way coupled drift-diffusion dynamics described in Section 2.5. Since the focus was on conducting a large set of parametric particle simulations, we further made the simplifying assumption of not modeling evaporation and particle size change. Individual occupants are assumed to not influence the airflow, and release respiratory particles in a one-way coupled manner representing respiratory exhalation, described in further detail in Section 2.6.

### 2.2 Geometry and Meshing Operations

3D CAD models of the single room for all six inflow-exhaust layout configurations were created using the cloud-based CAD platform OnShape [2]. For the door in closed configuration, an under-door clearance space of 0.075m × 0.915m was incorporated for infiltration airflow. Windows were similarly modeled using an opening of 0.025m × 1.25m for infiltration air flow. Inlet and outlet vents were modeled with an opening of 0.3m × 0.3m. In each model, flow extension regions were added at the locations of the two airflow vents, the door and the window each with an extension of 0.15m. Discretized mesh for the models were generated using built-in meshing packages in the cloud-based modeling suite SimScale [4]. Specifically, the hex-dominant meshing algorithm was employed with automatic mesh sizing, with a defined minimum edge length of 3.125 mm, and without boundary layer meshing. Local volumetric mesh refinement regions were added at the window and door entry zones, and the flow extension regions for the two vents. For all the simulations reported in the study, meshes with approximately 4.2 million cells (*4*.*6 million nodes*) were used.

### 2.3 Indoor Air Flow Quantification

Airflow through the room was computed by solving the governing equations for incompressible convective fluid flow based on the Boussinesq approximation for density variations using the SimScale suite, which utilizes the finite volume library OpenFOAM [3]. The steady state forms of the flow equations were solved for this study, to manage the overall computational load, and to evaluate the longer time scale averaged flow features that impact transmission risks. The governing equations used were the Reynolds Averaged Navier Stokes (RANS) fluid mass, momentum, and energy balance [20]. Effect of turbulence in the RANS framework was accounted for using the k-Omega Shear Stress Transport (SST) model. This choice is guided by the generally high ratio of accuracy vs computational expense, and superior performance in adverse pressure gradient boundary layers (*such as near floor or ceilings*). Velocity, temperature, turbulent kinetic energy, and turbulence dissipation rate were solved using the default smooth solver with Gauss-Siedel smoothing. Pressure was solved using the GAMG multigrid solver. The walls were given a temperature condition of 21.11^*◦*^*C* which was the same temperature used as the initial condition and temperature of the inlet, and no additional heating or cooling from air conditioning operations or presence of human occupants were modeled. The supply vent is modeled as a pressure inlet with zero gauge pressure. This is also employed for the entry regions in the door and the window. The exhaust boundary condition was set to 0.04 m^3^/s setting the ACH of the room to 3.0 which corresponds to am ASHRAE recommended value for a public office space. Relative residual tolerances for the solution variables were set at 1 × 10^−2^, and monitored for monotonic reduction of residuals (*final relative residuals for all cases are reported in supplementary material*). The final iteration data for the steady state flow variables were saved and post-processed for further analysis.

### 2.4 Indoor Flow Coherent Structure Identification

Coherent structures that organize mass transport in the flow were quantified using a Lagrangian computation of the Finite Time Lyapunov Exponent (FTLE) field based on the simulated airflow data. We employed a Cartesian grid-based tracer integration approach as demonstrated extensively in prior works [52,51]. A Cartesian grid of massless Lagrangian tracers were seeded in the flow domain, and their trajectories were computed by numerically integrating the flow velocity interpolated at the tracer locations as follows:

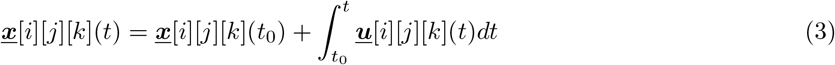

where [*i*][*j*][*k*] denotes the indexing scheme based on the Cartesian seed grid. Integration of the velocity requires efficient determination of the cell id where the particle resides at a given instant *t*. This was handled using a bounding level hierarchy based cell tree data structure [22] as implemented in the VTK library [5]. A deformation gradient 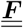 for tracer kinematics was computed for the mapping between the original configuration of the tracers at *t* = *t*_0_ (***<u>x</u>***[*i*][*j*][*k*](*t*_0_)) and the tracers at *t* = *t* (***<u>x</u>***[*i*][*j*][*k*](*t*)). as follows:

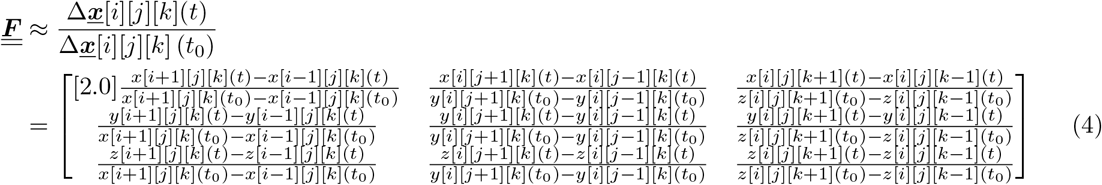

The corresponding right Cauchy Green deformation tensor was computed as follows:

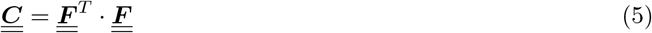

The FTLE field values were subsequently computed as a scaled version of the natural logarithm of the square root of the natural logarithm of the tensor 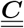 as follows:

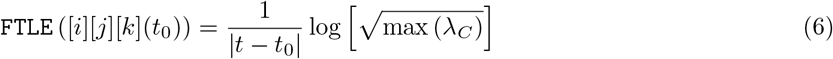

where *λ*_*C*_ denotes the eigenvalues of 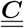. The FTLE notation on the lhs denotes that the computed FTLE field was mapped onto the original Cartesian grid used for seeding the tracers. This is the grid-based form of the computed FTLE field that was used for visualization of indoor air flow features. For this study we computed forward integration FTLE fields with a tracer grid resolution of 41 × 41 × 41 along the dimensions of the room (64, 000 *tracers*). All tracers were integrated for a period of 5 minutes.

### 2.5 Particle Transport Quantification

Pathogen loaded particles emanating from respiratory ejecta traverse the flow field based on local flow velocities and their own inertia. It is common to categorize these particles into large droplets, small droplets, and aerosolized droplet nuclei; and this size based distribution is connected to the type of respiratory event (*that is, normal breathing or forceful expiration such as coughing/sneezing*), mask wearing, and other factors. The effect of size and inertia is commonly quantified using the Stokes Number (*St*) - the ratio between particle momentum response time 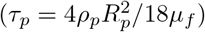 and a characteristic flow time scale (*τ*_*f*_). Here, we are particularly interested in a simplified model for fine-sized micro-particles, which are associated with small *τ*_*p*_, small *St*, and consequently their dynamics nearly follows the background flow. This dynamics is modeled using a simple drift-diffusion dynamics equation for the fine respiratory particles as follows:

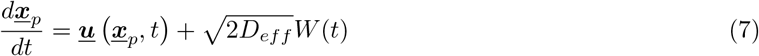

where the second term is a stochastic term representing the diffusion of particles in the flow. Following standard Brownian dynamics convention, this term is modeled as a scaled stochastic stationary Gaussian process with zero mean (*W* (*t*)). *D*_*eff*_ here represents the effective diffusion coefficient of the particles leading to the time scale of dispersion across a characteristic length *L*_*c*_ to 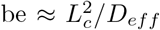. This includes diffusion due to Brownian motion as well as added dispersion induced by turbulence effects. As stated in Section 2.1, four cases with parametrically varied single effective diffusion coefficient *D*_*eff*_ = 0, 1e-6,, 1e-4, and 1e-2 m^2^/s were considered. The drift-diffusion equation is numerically solved using an explicit Euler-Maruyama scheme [7] with numerical time-step ∆*t* as follows:

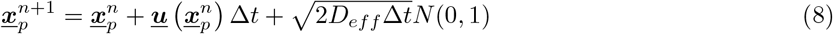

where *N* (0, 1) denotes a Gaussian random variable with zero mean and unit standard deviation. The resulting particle trajectories were computed using the same cell tree based particle location algorithm as in Section 2.5, and using absorbing boundary conditions at all walls.

### 2.6 Exposure Index Quantification

For each of the five infected index hosts, around 2700 particles were released in a spherically symmetric manner at their head location, which was assumed to be at a height of 5 feet 8 inches from the floor. The spherical release was employed as a simplification to aggregate various orientations in which the infected host may exhale. Sample size of around 2700 was chosen to ensure that a sufficiently large number of particles was released with a reasonable spatial density in a spherical surface, while still managing overall computational costs when considering multiple human occupants. For each infected host a total of 11 candidate subjects for viral exposure were considered, including the four other hosts. The locations defining the occupany vector **<u>Λ</u>**_*o*_ were selected such that: (a) they represent a range of inter-personal distancing (*including <* 3 *ft*., 3 − 6 *ft*., *and >* 6 *ft*.) to account for short-range and long-range transmission dynamics; and (b) they include varying combinations of subject location with respect to dominant FTLE ridges in the room (*more details provided in supplementary materials*). As particles reach the vicinity of the subject’s head, we evaluated the extent of exposure based on exposure zones modeled geometrically as spherically symmetric shells of thickness 2.5 cm each extending up to 10.0 cm from the head [48]. Four such zones were considered: Zone 1 included the head and 2.5 cm out from the head; Zone 2 included region between 2.5 cm and 5.0 cm from the head; Zone 3 included the region between 5.0 cm and 7.5 cm from the head; and Zone 4 included the region between 7.5 cm and 10.0 cm from the head (*see also details in supplementary material*). The Lagrangian exposure index *V E*_*L*_ was computed based on the particles entering and residing in these exposure zones, and a measure of their viability modeled based on viral half-life data from literature. Specifically, a history variable denoting elapsed time from instance of release is tracked for each particle, and its viability is modeled as an exponential decay function with specified half-life. Particle entry and exit into each spherical shell exposure zone is tracked, and weighted based on their distance from the head. Zone 1 is weighted at 100 %; zones 2, 3, and 4 weighted at 75, 50, and 25 % respectively. This results in the following form of exposure index for susceptible subject *s* from infected host *p*:

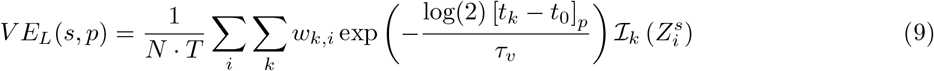

where *k* denotes index for particle; *i* denotes exposure zone index; 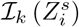 is an indicator function that has value= 1.0 if particle *k* is in the exposure zone 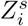 and zero otherwise; *w*_*k,i*_ denotes the weights described above; *τ*_*v*_ is the viral half-life parameter; [*t*_*k*_ − *t*_0_]*p* denotes the elapsed time-history for particle *k* released from infected host *p*; *N* is the total number of sample particles considered in the simulation; and *T* is the total duration of the particle tracking. For the purpose of this study for SARS-CoV-2, we have used the viral half-life data in aerosol mode as 4140 sec based on data in existing literature [56]. Note that, for interpretation of the index value, if all *N* biologically viable particles released by a host end up being at the face of another subject for entire time duration *T* the corresponding *V E*_*L*_ will be unity. Consequently, for all purposes, we expect *V E*_*L*_ value to be substantially lower than unity. Using the time-varying particle trajectory data obtained from the particle transport simulations as described in Section 2.5 the index *V E*_*L*_ was computed and interpreted in a pair-wise manner between infected hosts and susceptible subjects.

## 3 Results

### 3.1 Visualization of Indoor Flow with Varying Vent Positioning

Computed airflow patterns for each of the six model **<u>Λ</u>**_*v*_ configurations were visualized at three cross-sectional slices taken along the length, width, and the height of the room. Airflow patterns were visualized using the surface line integration convolution (LIC) maps on each slice. Surface LIC is a texture advection based technique for visualizing vector fields [13] that clearly demarcates the local rotational or vortical regions (*that is, vortex cores*) as well as the local flow velocity magnitudes (*based on LIC color mapping*). These illustrations are compiled in Figure 2 for all models where vents are located on opposite walls (*that is, 180UU, 180UL, 180LL configurations*), and in Figure 3 for all models where vents are located on orthogonal walls (*that is, 90UU, 90UL, and 90LL configurations*). Left panel for each row of Figures 2 and 3 denote sectional slice taken width-wise viewed from the wall with the door. Middle panel denotes sectional slice taken length-wise viewed from wall perpendicular to the door. Right panel denotes sectional slice taken midway along the height of the room viewed from the top. Maximum cell-based Reynolds numbers computed based on effective cell radius values were approximately in the range of 1,200 for all six cases. Slow flow velocities in the order of 0.1 m/s span the room interior with multiple prominent local circulatory flow regions influenced by the inflow and exhaust from the vent, and flow seepage due to the door and window (*see also sample animations in supplementary material*). The choice of temperature boundary conditions as stated in Section 2.3 leads to small temperature variations in the order of 1^*◦*^ C, with no high temperature gradients - a reasonable operating condition when no additional heating or cooling is modeled.

**Figure 2:**
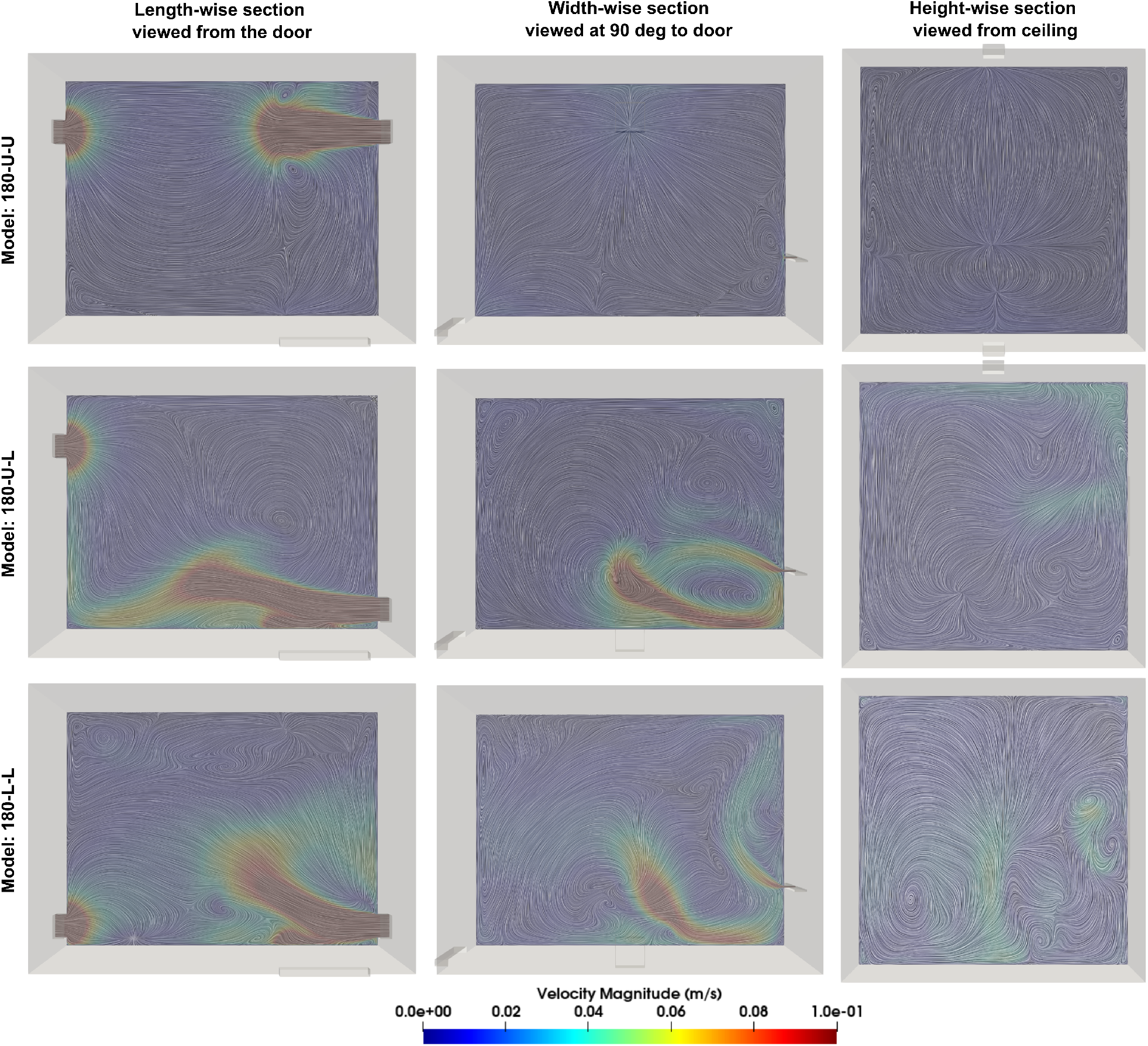
Visualization of computed air flow for rooms with vent-exhaust on opposite walls. Flow is viewed along sections cut across the length (left); width (middle); and height (right) respectively. See also sample animation provided with supplementary material.

**Figure 3:**
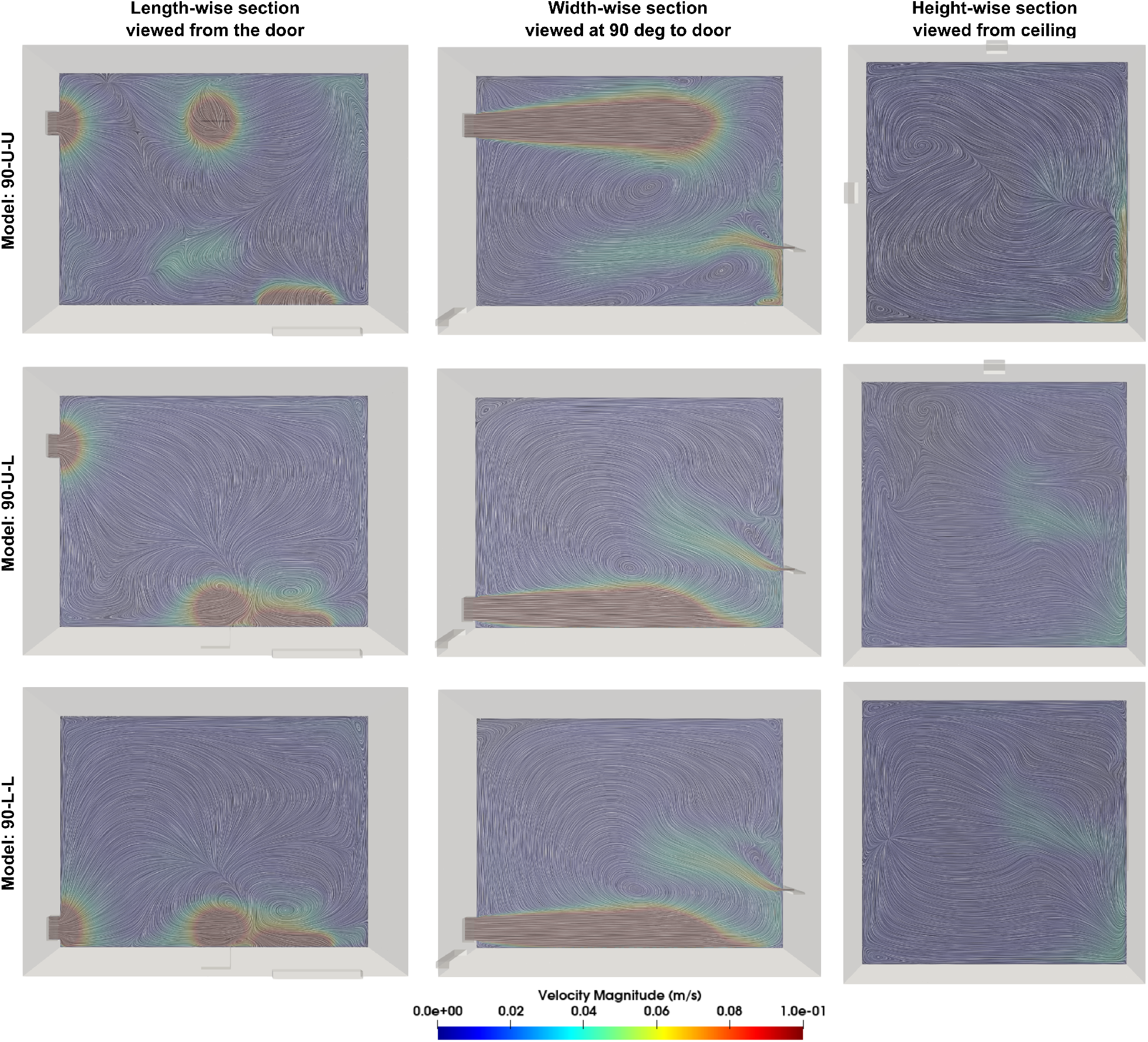
Visualization of computed air flow for rooms with vent-exhaust on perpendicular walls. Flow is viewed along sections cut across the length (left); width (middle); and height (right) respectively. See also sample animations provided with supplementary material.

The resulting flow patterns lead to the formation of local pockets and barriers organizing advective transport, whose boundaries can be identified by ridges in the computed FTLE field associated with the flow. For this study, forward time FTLE fields, which comprise inflowing manifolds for the flow, are utilized. Computed forward FTLE patterns in the interior of the room are compiled and illustrated in Figure 4. In this figure, the six room configurations are viewed along each row. Columns from left to right, for each room configuration, denotes FTLE fields viewed at sections from 0.75 m - 1.75 m in height in intervals of 0.25 m. Successive slices are selected to illustrate the contiguous structures that are formed by the FTLE ridges (*see also sample animations in supplementary material*). We clearly observe that prominent contiguous coherent structures originate from the ventilation driven indoor airflow pattern. These structures take complicated, non-uniform, curved and tortuous 3-dimensional shapes, and are not intuitive based on flow velocity field information alone. We note that these structures based on FTLE fields computed using RANS data indicate the averaged larger scale flow coherent structures, since finer scale flow feature information is inherently not resolved in RANS approaches.

**Figure 4:**
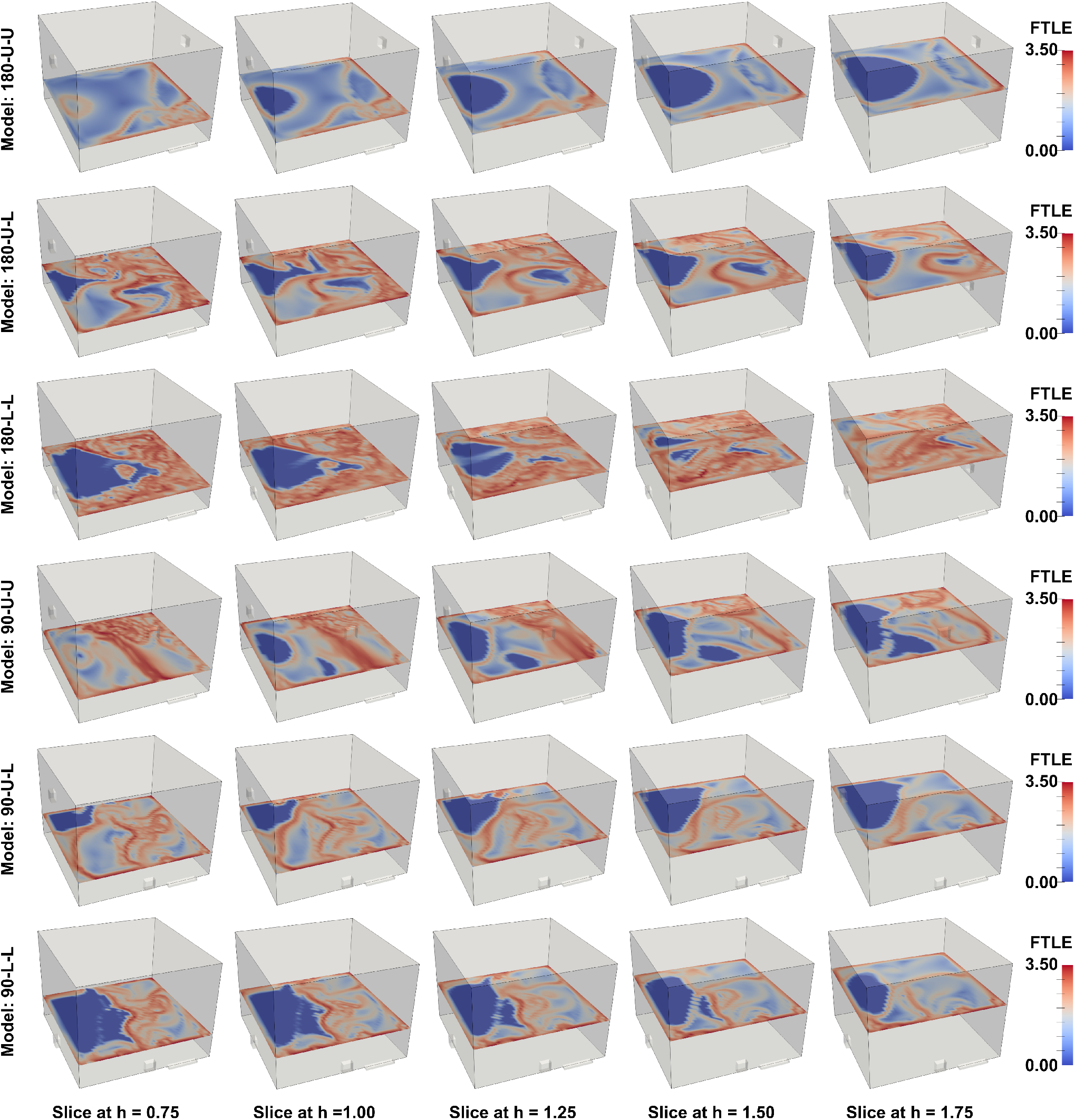
Computed forward FTLE field values shown across five successively varying sections taken along the height of the rooms. Each room is represented along the rows, and slice locations varied along the columns in this illustration. Ridges in FTLE would indicate locations of the coherent structure manifolds. See also sample animations provided with supplementary material.

### 3.2 Key Trends in Exposure Index Estimates

*V E*_*L*_ data was computed and compiled for all the 120 cases obtained by parametric variations in **<u>Λ</u>**_*b*_, **<u>Λ</u>**_*v*_, **<u>Λ</u>**_*p*_ and for all the 11 subjects for each of the five infected hosts (**<u>Λ</u>**_*o*_), leading to a total of 1,320 exposure indices quantified. The parametrically generated nature of this dataset was utilized to explore trends in exposure patterns, identify cases with low or high exposure, and investigate underlying flow physics phenomena as possible determinants for the same. For each host-subject pair, given their known locations as part of the occupancy vector **<u>Λ</u>**_*o*_, their physical separation distance was determined. Corresponding exposure indices for each of these separation distances are illustrated in a scatter plot for all four effective diffusivity values considered in Figure 5. The data indicates that (*in aerosol/micro-particle form as considered here*) there is both short-range transport and exposure over close distances (*less than 3 ft. or 0*.*9 m*) as well as long-range transport beyond the standard recommended distance of 6 ft or 1.83 m. This in turn indicates that, depending upon the ventilation and occupancy pattern in the room (**<u>Λ</u>**_*v*_ and **<u>Λ</u>**_*o*_), there may be chances of infection exposure even when physical distance of 6 ft or more are followed. This quantitative data further supports many expert recommendations on continued face mask usage and improved ventilation in indoor spaces despite distancing [8,24]. The exposure index dataset was further processed to isolate the effect of vent positioning (*variations in* **<u>Λ</u>**_*v*_). Specifically, slicing the dataset to extract all exposure data for each vent position, a sample set of 220 different exposure indices were extracted for each of the 6 models, and these samples are illustrated in Figure 6. The sample shading opacity is commensurate with the mean of the dataset - that is, higher mean indicates darker shading. Additionally, in Table 1 the maximum values, mean values, standard deviation, and the coefficient of variation (*ratio of standard deviation to the mean*) of these extracted sample sets are presented. It is observed, that for all the parameters (**<u>Λ</u>** *combination*) considered in this study, the 90LL configuration - that is, vents placed on orthogonal walls with exhaust towards the floor - leads to the lowest average exposure index, lowest standard deviation, as well as the lowest maximum value of exposure index. Conversely, the 90UU configuration - that is, vents placed on orthogonal walls with exhaust towards the ceiling - leads to the highest average exposure index, and standard deviation. The highest maximum value of exposure is seen with the 180UU configuration - vents placed on opposite walls with exhaust towards the ceiling. Finally, even with small scale temperature variations, the resulting particle distributions for convective flows when compared against those for flow without any thermal variations yields some notable differences, illustrating the need of accounting for proper thermal convection effects in indoor flows (*see related additional data presented in supplementary material*).

**Figure 5:**
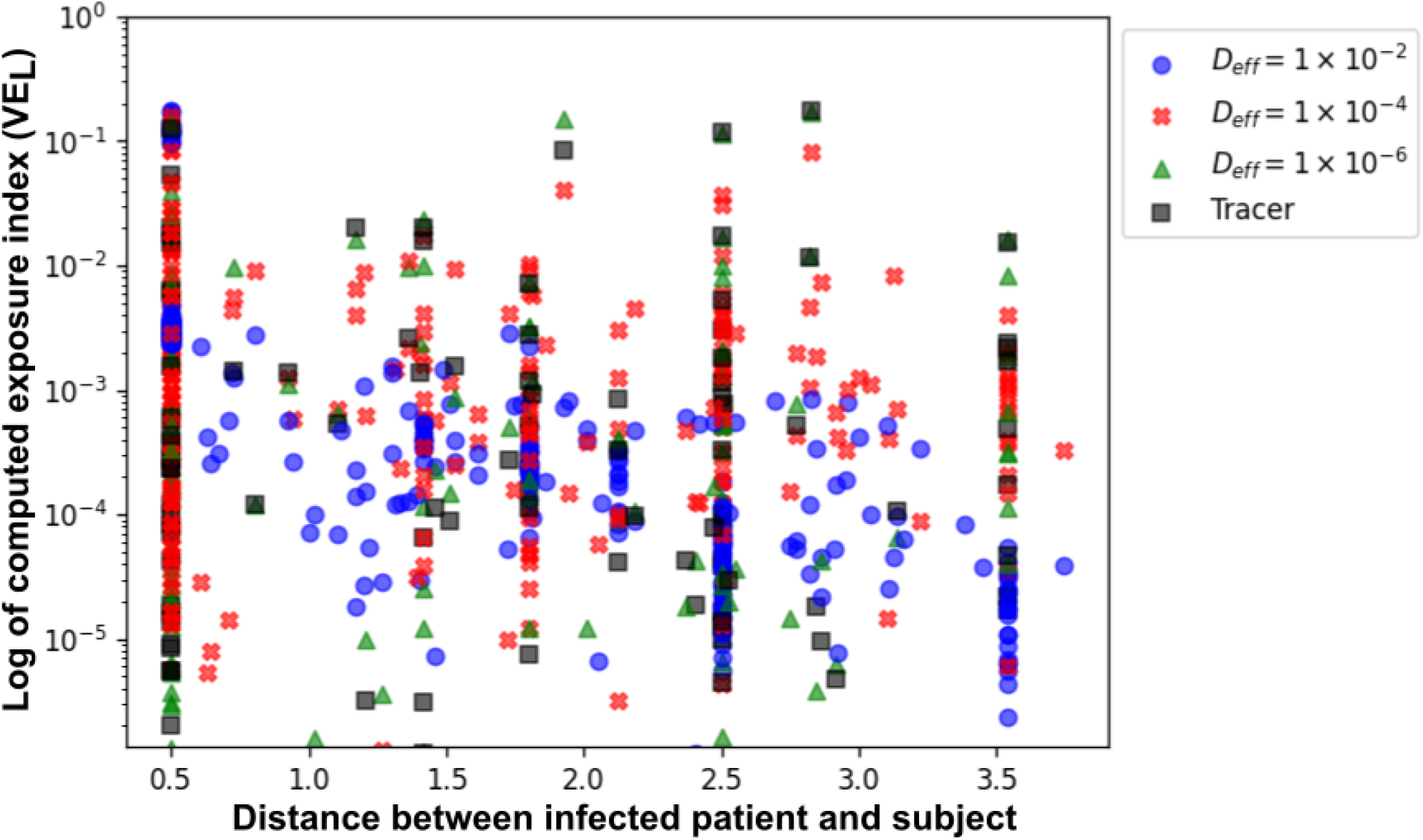
Summary of all 1,320 computed V E_L_ values for all cases considered in this study, presented as function of varying inter-personal physical distance. Distances are in meters (6 ft being at 1.83 meters approx.)

**Figure 6:**
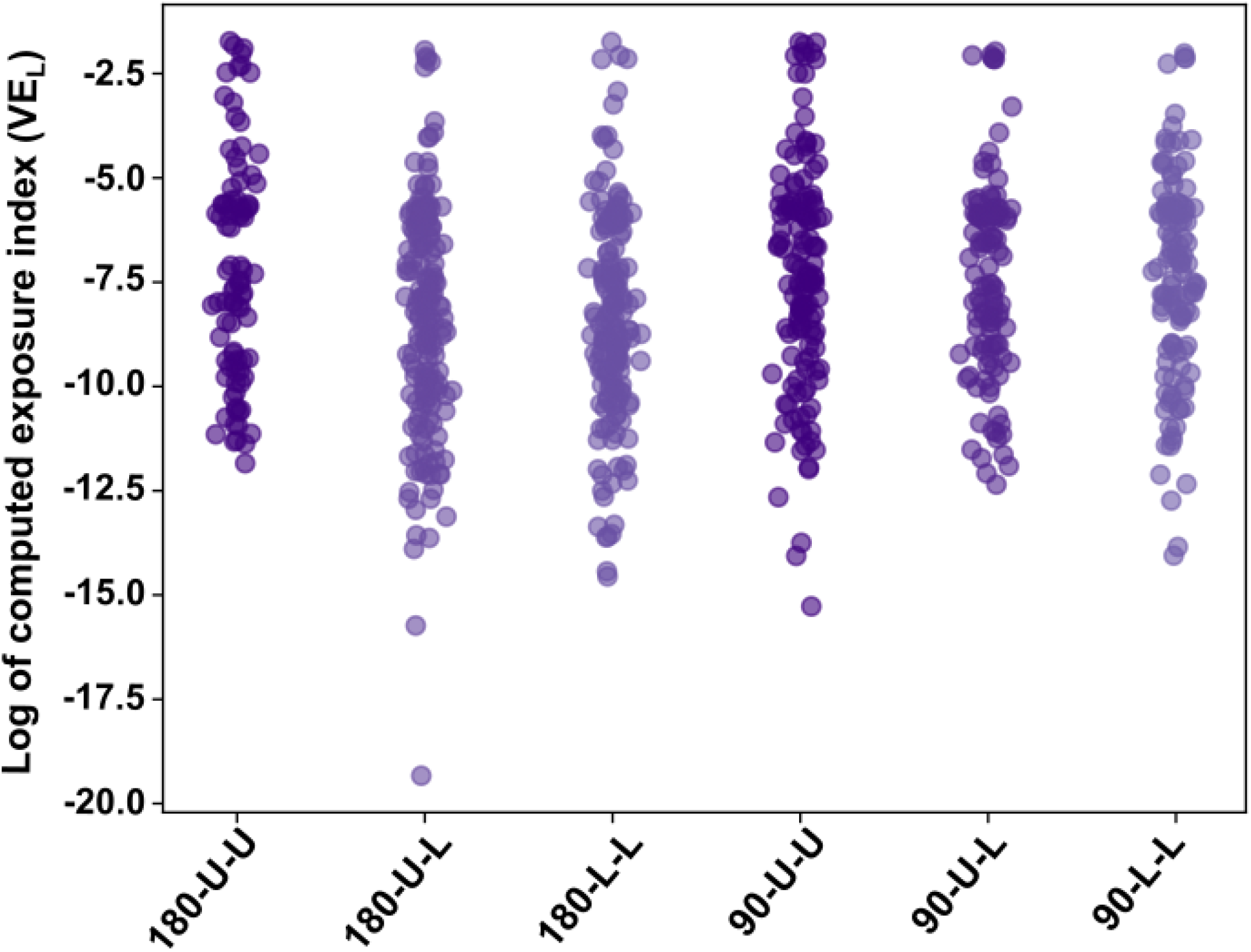
Illustration of computed V E_L_ samples categorized for each room across all parameters. Sample color shading is commensurate with mean values.

**Table 1:** Table presenting summary statistics of computed exposure index V E_L_ values categorized for each room. Entries in red denote the maximum, and those in blue denote the minimum for each statistic. The 90LL case clearly has the lowest mean V E_L_, max V E_L_, and standard deviation.

| Model | Max $VE_L$ | Mean $VE_L$ | Stdev $VE_L$ | C.O.V $VE_L$ |
| --- | --- | --- | --- | --- |
| 180UU | **0.1780** | 0.0058 | 0.0243 | 4.19 |
| 180UL | 0.1430 | 0.0039 | 0.0177 | 4.54 |
| 180LL | 0.1750 | 0.0038 | 0.0187 | 4.92 |
| 90UU | 0.1740 | **0.0076** | **0.0283** | 3.73 |
| 90UL | 0.1390 | 0.0044 | 0.0204 | 4.64 |
| 90LL | **0.1330** | **0.0036** | **0.0162** | 4.50 |

### 3.3 A Mechanism of Flow-mediated Organization of Particle Transport

The computed FTLE field data was processed further to visualize qualitative representations of the geometric manifolds in the flow that organize advective transport. Strictly in a mathematical sense, these manifolds are ridges in the FTLE fields, and require numerical algorithms based on the Hessian matrix of the FTLE values and the eigenvalues of the Hessian [6]. Owing to the computational complexity of managing the many simulation cases, and the high Cartesian grid resolution needed, such computational approaches were not employed in this study. Instead, isovolumes of high FTLE values in a certain range were selected - and using surface connectivity identification, the largest continuously connected surface geometry from this isovolume was visualized for a given room configuration. Pathlines from the various particle computations for the five infected hosts were also computed, and overlaid along with these surfaces. The resulting visualization was generated for various cases considered in this study, and used to illustrate the underlying flow-mediated organization of viral micro-particle transport. Here we present this visualization for a specific subset of those cases which lead to *V E*_*L*_ values of 0 for all subjects, including the ones placed in short-distance (0.5 m) from the infected host. The corresponding illustration is compiled in Figure 7. We observe that the resulting indoor air flow kinematics leads to the formation of non-intuitive pathways - which follow the FTLE field structures - that transport the particles away from subjects, and towards the exhaust vent. The pathlines for tracer particles follow these structure surfaces as shown in Figure!7, which is expected based on the definition of FTLE and associated coherent structure. We observe also that these pathlines for tracers with low effective diffusivity *D* =1e-6 m^2^/s follow the structure surfaces closely. This is likely due to the high effective Peclet number, in turn making the transport of these particles advection dominant.

**Figure 7:**
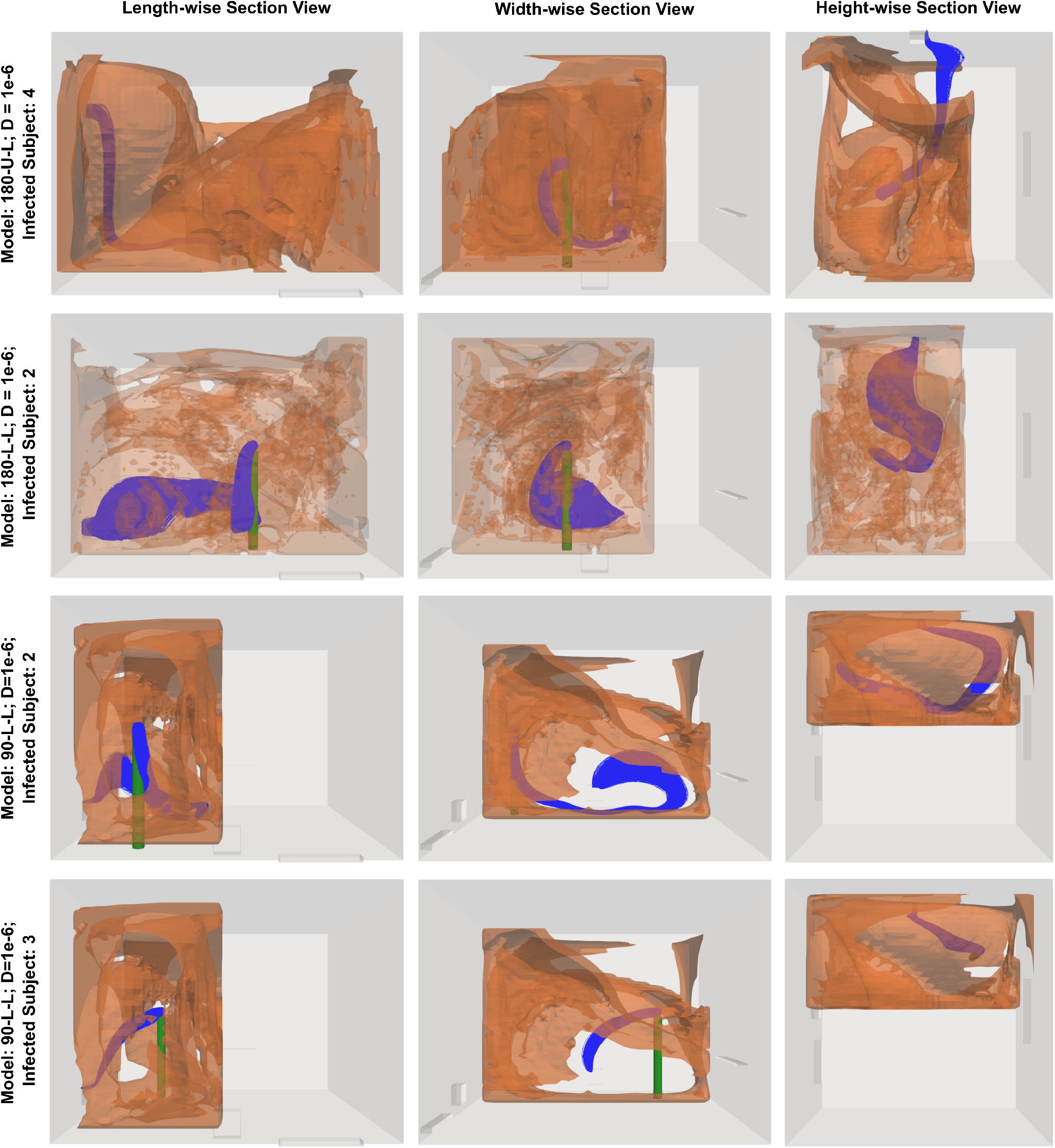
Illustration of a subset of four cases from amongst those leading to V E_L_ = 0, where extracted structures denoting FTLE isovolumes (in orange) are overlaid on computed pathlines (in blue). The FTLE structures are cut midway into the room to enable visualization of internal aspects.

## 4 Discussion

Here we have discussed two key contributions through our computational case-study: (a) the development of a parametric Lagrangian quantifier for viral/pathogen exposure in indoor spaces; and (b) the illustration of an underlying flow-mediated organization of viral/pathogen particle transport and exposure between occupants. For the former contribution, results from the numerical case studies conducted here establish the efficacy of our proposed parametric Lagrangian approach to compute and quantify a metric for determining infection exposure risk (*the exposure index V E*_*L*_). Rapid parametric computation of such quantifiers for infection transmission have broad implications. Our proposed framework can simultaneously account for major groups of parameters and their interplay - including space, layout, ventilation, occupancy, and pathogen related parameters. While the current study focused on methodology development, and on illustrating this framework on aerosolized micro-particles, our framework can be directly extended to study transport of larger pathogen-laden respiratory ejecta particles by accounting for appropriate physics of individual particles. This is an aspect of our ongoing and future work. The ability to compute quantifiers for infection exposure and risks parameterized against variables such as space, layout, ventilation, and occupancy can have large-scale implications and utility in informing decisions on indoor operations, ventilation modifications, and occupancy recommendations for schools, offices, and other businesses [39,40]. Additionally, while the current study mainly considers respiratory infections, with a focus on the SARS-CoV-2 owing to the recent global pandemic, pathogen transmission through airborne routes and their analysis in indoor spaces is also an important factor in several other infectious diseases. These include respiratory infections such as SARS, MERS, H1N1 etc. as well as other disease such as tuberculosis and measles [9,55]. By modifying the procedure to account for appropriate pathogen related variables (*e*.*g. inoculum size, viability, half-life, droplet/particle size etc*.) the framework can see broader applicability in such diseases as well.

Beyond the computation of an exposure index quantifier, the flow physics based analysis of exposure index values and the associated FTLE based Lagrangian analysis conducted here reveal the complex non-intuitive spatio-temporal pathways for respiratory micro-particle transport in indoor spaces. We have illustrated here that these otherwise hidden pathways have a potentially key role to play in determining the extent of long-range as well as short-range transmission dynamics. Specifically, for some of the cases with 0 exposure index values, we illustrated this in Section 3.3. The data and analysis in Section 3.3 clearly show that the embedded manifolds highlighted by the FTLE fields can shape the transmission pathway by routing respiratory particles within the indoor space. We remark that the usage of Lagrangian analysis of coherent structures and FTLE in indoor infection transmission context has been sparsely investigated, and this study establishes the utility and efficacy of this technique for micro-particle cases. When extended to larger respiratory particles, the technique still holds applicability. We have considered in this study a drift-diffusion dynamics, and interpreted the corresponding advection dominant (*high Peclet number*) dynamics using the FTLE. For larger particles, considering particle inertia, an alternative inertial particle FTLE can be computed as proposed in several other works [46,53]. Inclusion of stochastic components in the dynamics will not deviate substantially from FTLE based intuition of transport so long as the associated Peclet number is high (*that is, the transport is advection dominant*) [25]. This is also illustrated in Figure 7, where a low but non-zero effective diffusivity particle transport is shown to follow FTLE structure boundaries. Considerations of particle size and inertia are also of particular relevance when argued based on the particle momentum response time, defined for spherical particles in Section 2.5. Conceptually, for a single particle with a momentum response time *τ*_*p*_, the velocity of that particle within a constant flow of *u* starting from 0 under low Reynolds number environments is:

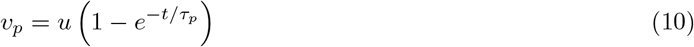

Assuming respiratory ejecta particles in the range 100 microns or lesser, the estimated response time is 30 ms or lesser. Hence, within an extremely short time, the particle will respond appreciably to take up local flow velocity - thereby remaining possibly entrained and suspended, for sufficiently large durations, within local circulatory regions of flow - which in turn will shape the manifolds highlighted by the FTLE fields.

Several additional incorporations can be made to the model presented for this study, potentially aligned with the model assumptions stated in Section 2.1. While a steady state flow assumption was made for the cases considered here, the framework can be applied as proposed also for unsteady flow data. Similarly, as discussed above, despite only considering micro-particle cases here, the framework can be applied the same way for larger scale particles or droplets. Furthermore, while the current treatment of a single room was simplified (for managing the scope of this 120 simulation case study), more complicated room layouts including furniture and other larger installations can be incorporated easily into the room geometry. While the inclusion of these various levels of detail will add accuracy to the model and its predictions, they will also incur added computational costs. One key limitation for the cases considered in this study is that the exact human geometry and associated thermal convective boundary layer [31] was not incorporated. Our approach also does not currently account for whether an individual host is a super-spreader, whether an individual subject is a more susceptible individual, or whether any subject is wearing a mask. Additionally, while aspect of biological viability of the particles considered here has been included, the actual number of virions inside each such particle may be higher and may differ depending upon the pathogen and the disease. Incorporation of these details are ongoing efforts towards expanding the model capabilities and fidelity.

## 5 Concluding Remarks

Here we have presented a Lagrangian computational framework towards computing a quantifier for the extent of human-to-human viral/pathogen exposure in indoor spaces - referred to as the Lagrangian Viral Exposure index (*V E*_*L*_). The mathematical and algorithmic foundations of this computational framework are outlined. We conduct a systematic study on a simplified room configuration to illustrate the flow mediated transport pathways and mechanisms for viral particle transport using the concept of Lagrangian Coherent Structures based on computed Finite Time Lyapunov Exponent (FTLE) fields for the indoor airflow. The results from a cohort of 120 simulations obtained from systematic variations of key parameters demonstrate the efficacy of the proposed approach, and highlight key determinants of infectious micro-particle transport between human subjects in an indoor space, and the role of complex flow-mediated organization of micro-particle transport governing long-range and short-range transmission.

## Supporting information

Flow field visualization for the 90UU case referred in manuscript.

FTLE field visualization for the 90UU case referred in manuscript.

Flow field visualization for the 180UU case referred in manuscript.

FTLE field visualization for the 180UU case referred in manuscript.

## Data Availability

All associated data have been presented in the manuscript.

## 7 Acknowledgements

This work utilized resources from the University of Colorado Boulder Research Computing Group, which is supported by the National Science Foundation (awards ACI-1532235 and ACI-1532236), the University of Colorado Boulder, and Colorado State University. The Authors also acknowledge the availability of an academic license from SimScale to complete this work. DM designed the study and conducted FTLE computational analysis; JW designed all models, parametric simulations, computational fluid dynamics, and particle transport computations; SM co-designed data analysis and interpretation in the context of infection transmission, and guided parameter selection. All authors have reviewed and agreed to the final draft of the manuscript.

## Supplementary information

### Animation of Flow and FTLE data

A set of four animations have been included along with the manuscript. Two animations - named *180UU-FlowAnimation*.*mov* and *90UU-FlowAnimation*.*mov* - illustrate the computed flow patterns. The other two - named *180UU-FTLEAnimation*.*mov* and *90UU-FTLEAnimation*.*mov* - illustrate the computed forward FTLE field for the same two rooms for which flow animations are provided. These animations are provided to support the visualization of flow velocity patterns as computed and described in Section 3.1.

### Proof of Solution Convergence for all CFD Cases

For the RANS CFD model used to compute the steady state air flow patterns in all the 6 different room configurations, all solution field variables were monitored for convergence such that the reported relative residuals from the underlying finite volume solver in SimScale for all solution variables were below 1 × 10^−2^ (the relative residual tolerance). All solutions presented here satisfied this convergence criteria based on relative residuals. Here, we have compiled the final relative residual values for all solution variables and for all cases to support our claim of convergence.

**Table S1:** Table presenting reported relative residual values for final iteration of the steady flow simulations presented here. All variables had a relative residual tolerance of 1e-2.

| Case | Ux | Uy | Uz | k | $\omega$ | p (rel.) | temp. |
| --- | --- | --- | --- | --- | --- | --- | --- |
| 90LL | 2.77e-04 | 1.31e-03 | 1.19e-03 | 5.32e-05 | 1.92e-06 | 5.60e-03 | 1.58e-03 |
| 90UL | 2.84e-04 | 1.33e-03 | 1.24e-03 | 5.37e-05 | 1.96e-06 | 6.15e-03 | 1.54e-03 |
| 90UU | 2.45e-04 | 1.06e-03 | 1.13e-03 | 5.43e-04 | 1.27e-06 | 3.01e-03 | 1.67e-03 |
| 180LL | 2.90e-04 | 7.61e-04 | 1.00e-03 | 6.84e-04 | 1.92e-06 | 3.18e-03 | 2.92e-03 |
| 180UL | 1.92e-04 | 5.30e-04 | 8.52e-04 | 3.45e-04 | 1.19e-06 | 2.50e-03 | 1.32e-03 |
| 180UU | 3.31e-04 | 6.37e-04 | 1.64e-03 | 7.42e-04 | 5.91e-07 | 9.62e-03 | 8.41e-05 |

### Details on Exposure Zones

Figure S1 provides a visualization of a portion of the exposure zones as discussed in Section 2.5. In this representation only a section of exposure zone is shown for ease of viewing. For the calculations the exposure zones were spheres that enclosed the head. The weighted exposure zones used in the *V E*_*L*_ calculation were set up to have a linearly spaced radius between the surface of the head and 10 cm out from the surface of the head. In this calculation, four exposure zones were used so each zone had a radius that was 2.5 cm greater than the immediate interior zone.

### Subject Placement Details

Figure S2 shows the placement of the human subjects for each room configuration. Subjects 0 through 4 were iteratively selected as the infected host and these subjects are shown in red and yellow. Subject 0 is shown in red because in each configuration this setup is based on Subject 0 being the infected host. The four yellow subjects are the other iterative subjects. Subjects 5 through 8 are shown in orange to distinguish them as the nearby subjects. These subjects move with the infected host and their positions are based relative to the infected host; i.e. Subject 5 is always 0.5 m in the +y direction from the infected host. Subjects 9 through 11 were placed based on an evaluation of the FTLE fields of the flow for that configuration and are shown in purple.

**Figure S1:**
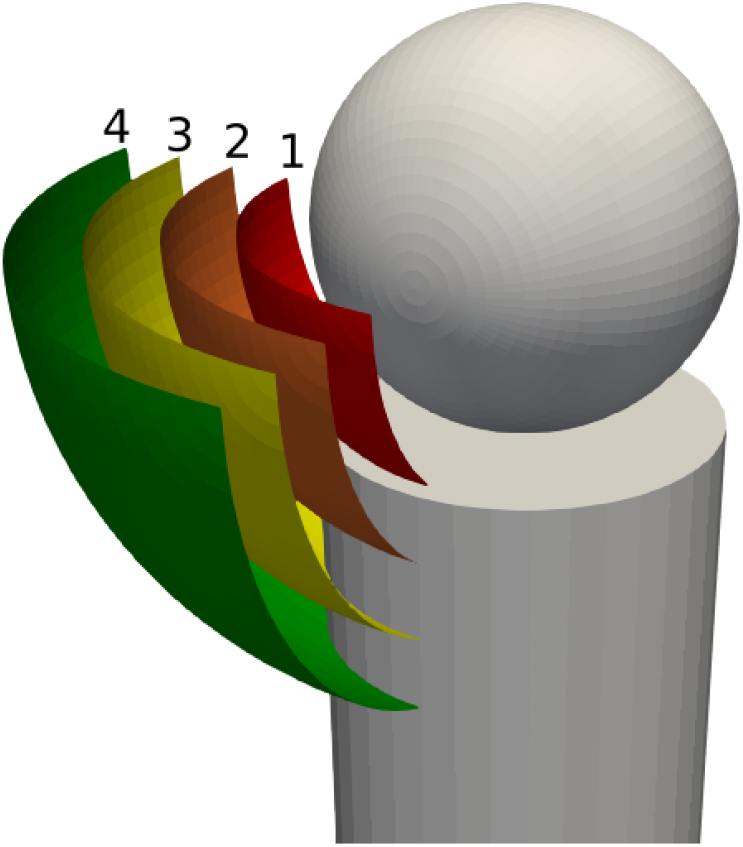
Visualization of Exposure Zones around a subject for computation of V E_L_. The gray sphere and cylinder show the position of the subject head.

### Comparison with Non-Convective Case

The thermal effects in an enclosed space can greatly influence the exposure calculations. Even in cases where temperature fluctuations are small, such as the data presented in this paper, the effects are noticeable and measurable. Comparing Table 1 with Table S1 shows that for the convective cases, the configuration with the lowest possible exposure risk was clearly the 90-L-L configuration whereas for the non-convective case the corresponding configuration was the 180-U-L case. For a comparison across all configurations between convective and non-convective cases, Figure 6 can be compared with Figure S3 below. Both figures present the shading on the scatter data as a function of the mean; the higher the mean, the more opaque the points. Through this comparison it can be easily seen that the 180 cases consistently present a lower mean exposure index, and this is more evident in Figure S3 representing the non-convective cases.

**Figure S2:**
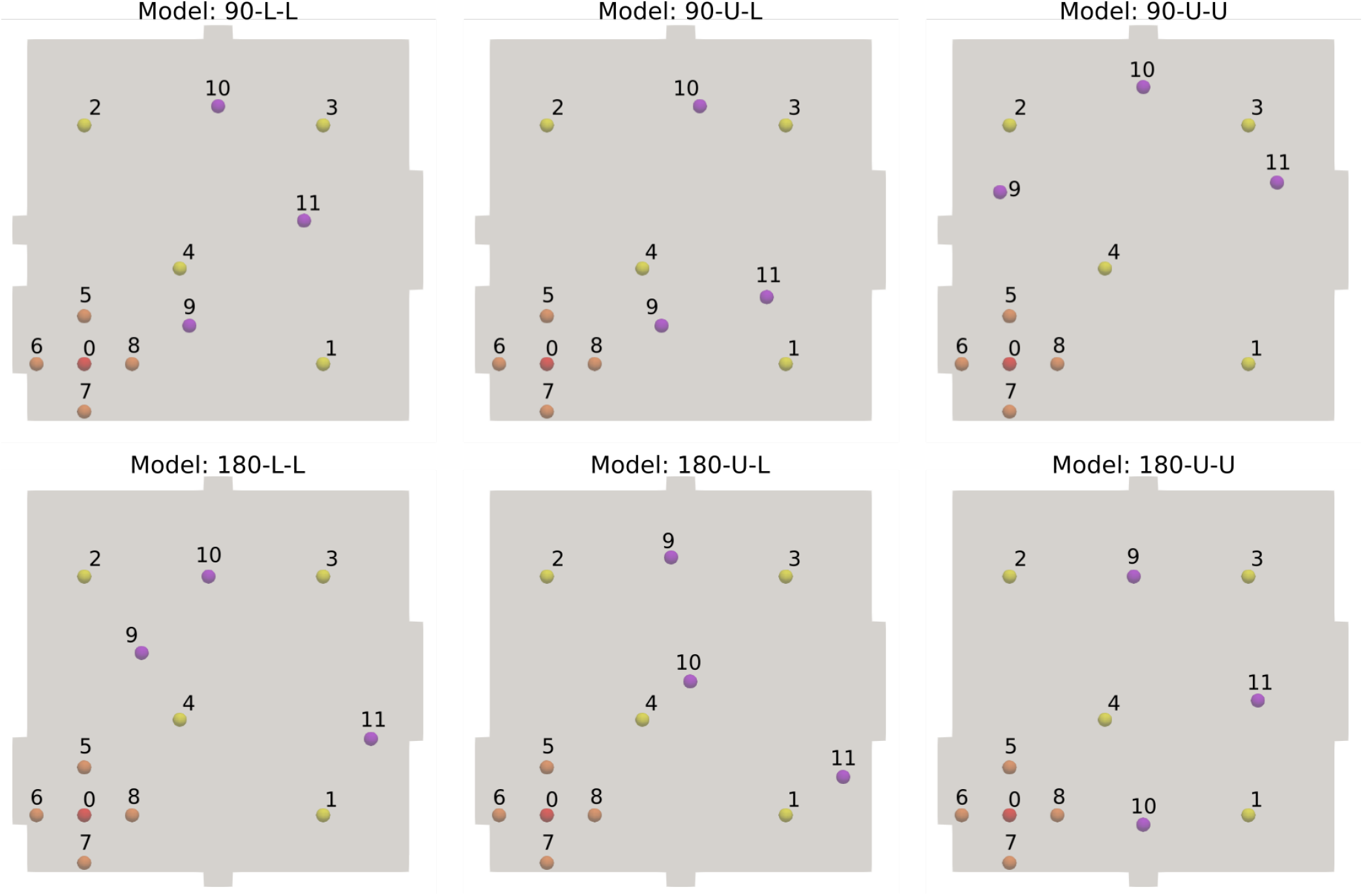
Placement of patients for each configuration when subject is Patient 0

**Table S2:** Summary statistics of the computed V E_L_ for the non-convective runs.

| Model | Max $VE_L$ | Mean $VE_L$ | Stdev $VE_L$ |
| --- | --- | --- | --- |
| 180UU | **0.1640** | 0.0051 | 0.0215 |
| 180UL | **0.1240** | **0.0034** | **0.0150** |
| 180LL | 0.1320 | 0.0039 | 0.0173 |
| 90UU | 0.1410 | **0.0080** | **0.0231** |
| 90UL | 0.1550 | 0.0049 | 0.0191 |
| 90LL | 0.1500 | 0.0048 | 0.0183 |

**Figure S3:**
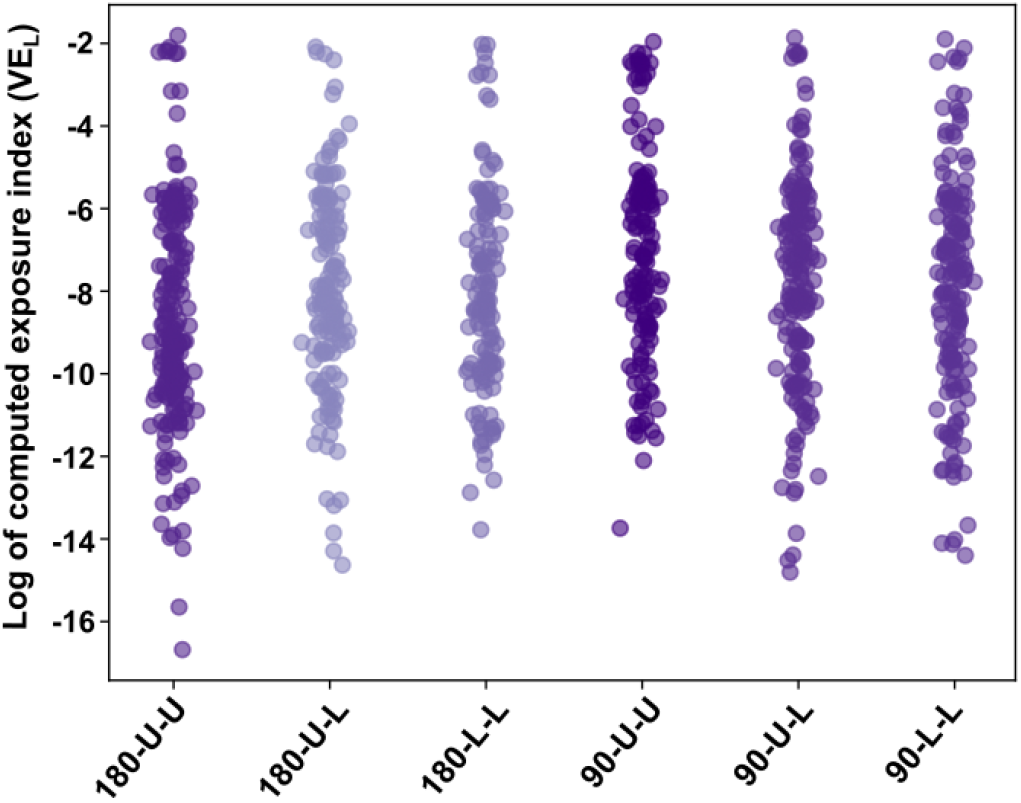
Scatter plot of the various exposure index samples categorized based on the rooms, for all parameter combinations, obtained from the non-convective runs.

